# Leveraging multimodal neuroimaging and GWAS for identifying modality-level causal pathways to Alzheimer’s disease

**DOI:** 10.1101/2025.02.27.25322897

**Authors:** Yuan Tian, Daniel Felsky, Jessica Gronsbell, Jun Young Park

## Abstract

The UK Biobank study has produced thousands of brain imaging-driven phenotypes (IDPs) collected from more than 40,000 genotyped individuals so far, facilitating the investigation of genetic and imaging biomarkers for brain disorders. Motivated by efforts in genetics to integrate gene expression levels with genome-wide association studies (GWASs), recent methods in imaging genetics adopted an instrumental variable (IV) approach to identify causal IDPs for brain disorders. However, several methodological challenges arise with existing methods in achieving causality in imaging genetics, including horizontal pleiotropy and high dimensionality of candidate IVs. In this work, we propose testing the causality of each brain modality (i.e., structural, functional, and diffusion MRI) for each gene as a useful alternative, which offers an enhanced understanding of the roles of genetic variants and imaging features on behavior by controlling for the pleiotropic effects of IDPs from other imaging modalities. We demonstrate the utility of the proposed method by using Alzheimer’s GWAS data from the UK Biobank and the International Genomics of Alzheimer’s Project (IGAP) study. Our method is implemented using summary statistics, which is available on GitHub.

## 1. Introduction

Studies indicate that dementia, including Alzheimer’s disease (AD), is highly heritable, and genetic factors are estimated to play a role in around 80% of AD cases (Gatz et al. 2006; Tanzi 2012). However, research on dementia still has significant room for developing genetics-brain pathways. Although genome-wide association studies (GWASs) have identified a number of genetic risk factors for AD, they are limited in testing for a direct association between the variants and the disease trait. GWASs do not provide a comprehensive understanding of how the genetically-regulated structural and functional brain pathways drive AD progression, which is essential for characterizing the genetic mechanism of the disease.

The growing interest in the role of genetics in brain disorders has spawned a new field known as imaging genetics that integrates neuroimaging features (e.g., from brain magnetic resonance imaging (MRI)) with genetics using statistics and machine learning. Many neuroimaging features, including the brain’s anatomy (structural MRI (sMRI)), function (functional MRI (fMRI)), and microstructure (diffusion MRI (dMRI)), have been demonstrated to be heritable. For example, Lin et al. (2014) and Huang et al. (2015) use imaging features as GWASs traits to identify the associations between the imaging features and single nucleotide polymorphisms (SNPs) or a set of SNPs (e.g., a gene). At the same time, in recent years, there has been substantial evidence in the studies of brain disorders that brain MRI features, i.e., imaging-driven phenotypes (IDPs), are useful biomarkers for AD and are modulated by genetic factors (Elliott et al. 2018). By integrating data from imaging scans and GWASs, researchers have attempted to better understand the heritability of AD and other brain-related diseases. Such integrative analyses have been promising in advancing imaging neuroscience that paces with an increased size of genetic and neuroimaging data (Shen and Thompson 2019). However, due to its high dimensionality (e.g., thousands of IDPs available from UK Biobank (Elliott et al. 2018; Smith et al. 2021)) and complex correlation structure (e.g., correlations between imaging features), it is difficult to fully leverage brain imaging data in GWASs, especially in pinpointing causal imaging features affecting the heritability of brain-related diseases (Burgess et al. 2017).

The Transcriptome-wide association study (TWAS), which leverages expression quantitative trait loci (eQTL) with GWAS, has become an important statistical tool in genetics research (Gamazon et al. 2015). TWAS identifies genes associated with diseases/phenotypes through genetic regulation of gene expression. TWAS is constructed by first using SNPs in a gene to predict the corresponding gene expression levels via machine learning methods (e.g., penalized regression), which represents the “genetically regulated” (or “genetically imputed”/”genetically predicted”) component of gene expression. The predicted gene expression levels are then associated with selected outcomes, adjusting for multiple comparisons. In neuroimaging literature, the imaging-wide association study (IWAS) extended TWAS by considering multiple IDPs as intermediate phenotypes rather than gene expression, which may offer improved mechanistic interpretation when studying brain disorders such as AD (Xu et al. 2017). These approches showed significant improvement in localizing genes associated with the phenotype compared to GWAS.

In addition, TWAS offers a statistical interpretation from the lens of causal inference via instrumental variable analysis. Specifically, TWAS is equivalent to testing a causal relationship from eQTL to phenotype using 2-stage least squares (2SLS) by using SNPs as instrumental variables (Xue et al. 2020; Mai et al. 2023). In neuroimaging, BrainXcan takes this approach to identify IDPs leading psychiatric traits under the assumptions of Mendelian Randomization (MR) (Liang et al. 2022). The multivariate IWAS (MV-IWAS) extends the causal interpretation of TWAS in imaging genetics by accounting for horizontal pleiotropic effects of other IDPs (Knutson et al. 2020). We refer to Taschler et al. (2022) for a comprehensive review of conducting causal inference with neuroimaging data using MR. A key consideration in these approaches in imaging genetics is the high dimensionality of the IDPs. For example, the UK Biobank provides thousands of IDPs, which may increase dramatically based on preprocessing pipelines and analysis goals (e.g., ROI-level analysis versus whole-brain analysis). Because it increases the burden of multiple comparisons, BrainXcan and MV-IWAS consider the whole genome as potential instrumental variables and use polygenic scores (PGSs) to implement the first stage of 2SLS. However, such approaches would lose the original purpose of TWAS, which was to localize gene-level associations with diseases/phenotypes.

This paper aims to mitigate current challenges in making causal interpretations in imaging genetics and localizing corresponding genes. Rather than evaluating each IDP individually, we propose to make causal interpretations between diseases/phenotypes and each *imaging modality* (e.g., sMRI, fMRI, or dMRI). Each modality represents distinct aspects of brain structure and function, and therefore, aggregating the brain information in each modality can contribute to a more comprehensive understanding of the genetic mechanisms underlying brain disorders. At the same time, our method explicitly accounts for horizontal pleiotropy among imaging modalities, therefore providing robustness of the causal direction between each modality and the progression of AD. With the MRI modalities through which genetics affect brain function in AD, we can better understand the underlying biological mechanism by which genetics causes changes in brain function, which in turn causes AD. We also make our methods implementable using GWAS summary statistics and reference panels, enhancing the practical utility.

The rest of the paper is organized as follows. In Section 2, we provide a methodological review of the statistical methods in imaging genetics that extended TWAS and develop the proposed method, including the implementation with summary statistics. In Section 3, we evaluate Type 1 error rate and power of the proposed method by simulations. Section 4 applies our methods to the AD GWAS data obtained by the UK Biobank and the International Genomics of Alzheimer’s Project (IGAP). A few points of discussion are made in Section 5.

## 2 Methods

### 2.1 Notations

We let **y** = (*y*_1_, …, *y*_*N*_)^*′*^ denote the phenotype of our interest (e.g., AD status) from *N* subjects. Also, let **g**_*j*_ = (*g*_1*j*_, …, *g*_*Nj*_)^*′*^ be the collection of the *j*th SNP across subjects in a gene for *j* = 1, …, *J*. Similarly, we let **m**_*k*_ = (*m*_1*k*_, …, *m*_*Nk*_)^*′*^ be the *k*th IDP for *k* = 1, …, *K*. Without loss of generality, we assume that each **g**_*j*_ and **m**_*k*_ are standardized to have the zero mean and unit variance.

### 2.2 Review of existing methods

We first review transcriptome-wide association study (TWAS) and relevant approaches in neuroimaging.

#### 2.2.1 TWAS and UV-IWAS

With a single endophenotype **m**, TWAS is a method that integrates endophenotypes (e.g., gene expression) in gene-based association study to test the genetically-regulated effect of the endophenotype on the disease trait (Gusev et al. 2016). TWAS is conducted using a two-stage approach. In Stage 1, using a set of SNPs encoded in a gene (**g**_1_, …, **g**_*J*_), machine learning methods (e.g., linear models) are used to predict a corresponding gene expression level (**m**) based on the following model:

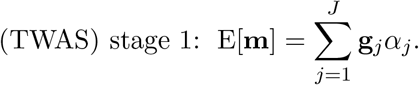

Here, 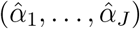 is obtained by fitting a penalized linear regression with the ridge (Liang et al. 2022), LASSO or elastic net penalty (Gusev et al. 2016). Then, a *genetically imputed* expression, 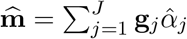 is used to test for its association with the phenotype (**y**) in Stage 2 working model:

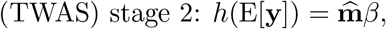

where *h*(*·*) is the canonical link function (e.g., the logistic link for binary traits and the identity link for for quantitative traits). TWAS tests *H*_0_ : *β* = 0 to identify genes whose genetically-regulated gene expression is associated to the trait. UV-IWAS is conceptually equivalent to TWAS (Knutson et al. 2020), using an imaging-driven phenotype (IDP) instead for gene expression as an endophenotype, implying that IDPs would be more useful than gene expression in finding associations between genotypes and brain disorders.

TWAS (or UV-IWAS) provides a useful framework that bridges the gap between genetic associations and molecular mechanisms, and it offers several possible statistical interpretations. One interpretation is that it is essentially a weighted burden test for gene-based association testing where the weight for the *j*th SNP in a gene is determined by 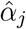 in Stage 1. Because an optimal choice of weights would lead to improved power in the burden test, the choice of weights in TWAS, which is informed biologically, expected to improve statistical power. Another interpretation is that it can be interpreted as an instrumental variable (IV) approach, where the 2-stage least squares (2SLS) is used to estimate and infer the *causal* effect of endophenotypes (**m**) on the phenotype (**y**). In such a case, the SNPs that led the causal direction serve as composite IVs, provided that the following assumptions hold.

A1. There exist correlations between all SNPS selected in Stage 1 and the endophenotype;

A2. SNPs are uncorrelated with potential confounders;

A3. SNPs do not affect the disease trait, except through the endophenotype/IDP being tested.

A3 indicates that the SNPs do not affect the disease trait via genetic direct effect. Since most SNPs are unrelated to endophenotypes, certain thresholds are typically used in TWAS Stage 1, such as predictive *R*^2^ or *p* value, to ensure that A1 is met.

#### 2.2.2 Multivariate IWAS (MV-IWAS)

A unique challenge in imaging genetics is that there are multiple IDPs each may have distinct genetic regulatory pathways and potential pleiotropic effects to AD. However, multiple significant IDPs have been identified (Xu et al. 2017), as discussed by Knutson et al. (2020) and Elliott et al. (2018), the presence of horizontal pleiotropy can lead to violations of the IV assumptions, resulting in invalid causal inference using univariate IV-analysis and ultimately inflating Type I error in the hypothesis testing for causal effects from an IDP to a phenotype, especially when there is high correlation between the IDPs.

To address the potentially inflated Type I error rate due to pleiotropy, Knutson et al. (2020) proposed using multiple linear regression in Stage 2 to adjust for possible pleiotropic effects of genetically-regulated IDPs from gene-to-disease. If there are *K* imaging features in total, MV-IWAS considers the working model the second stage:

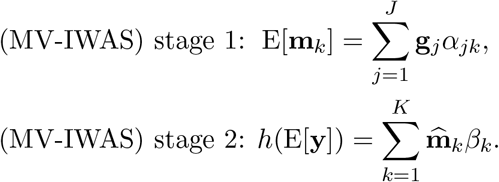

Stage 1 of MV-IWAS is equivalent to Stage 1 of TWAS/UV-IWAS, where each IDP is predicted separately by the same set of SNPs in a gene. In Stage 2, a general linear model (GLM) is applied using all predicted endophenotypes, replacing the univariate regression in TWAS/UV-IWAS. In MV-IWAS, whether the *k*th IDP causes the phenotype is achieved by testing *H*_0*k*_ : *β*_*k*_ = 0 in the Stage 2 model, which controls the pleiotropic effects of the other IDPs. Furthermore, to account for remaining genetic direct effects, Knutson et al. (2020) extended the Stage 2 model to MV-IWAS-Egger to account for direct genetic effects on the phenotype. The Stage 2 model of MV-IWAS-Egger is be formulated by

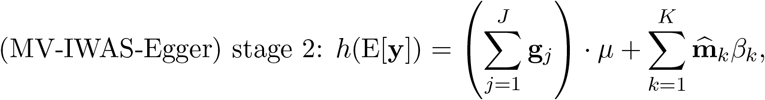

with the same null hypothesis *H*_0*k*_ : *β*_*k*_ = 0 to test the causal effect of the *k*th IDP on the phenotype. We note that, from its construction, the power of MV-IWAS and MV-IWAS-Egger is severely affected by the multicolinnearity of the 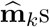(and 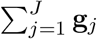 for MV-IWAS-Egger) (Burgess et al. 2017).

#### 2.2.3 Remaining challenges

While MV-IWAS provides a causal interpretation, it could be underpowered due to the following factors:

##### 1. Multicollinearity

Multicollinearity arises when two genetically regulated components of the IDPs (e.g., 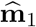 and 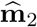) are highly correlated, leading to a challenge in point estimation (i.e., an increase in the standard error) in Stage 2 of MV-IWAS and a decrease in statistical power. We refer to Chan et al. (2024) for the challenges with applying Mendellian randomization in case of high dimensional exposures (IDPs).

We empirically evaluated the multicollinearity by computing the genetic correlations of the IDPs. Specifically, we performed Linkage Disequilibrium Score Regression (LDSC) analyses (Duncan et al. 2018) using UKB IDP GWAS summary statistics available from Smith et al. (2021). For computational reasons, we included top 50 features with highest heritability estimates from different imaging modalities. As shown in Figure 1, 28.5% of sMRI and 60.3% of dMRI within modality IDP pairs show genetic correlation greater than 0.7, which implies that including such highly correlated IDPs in MV-IWAS could suffer from power loss.

**Figure 1.**
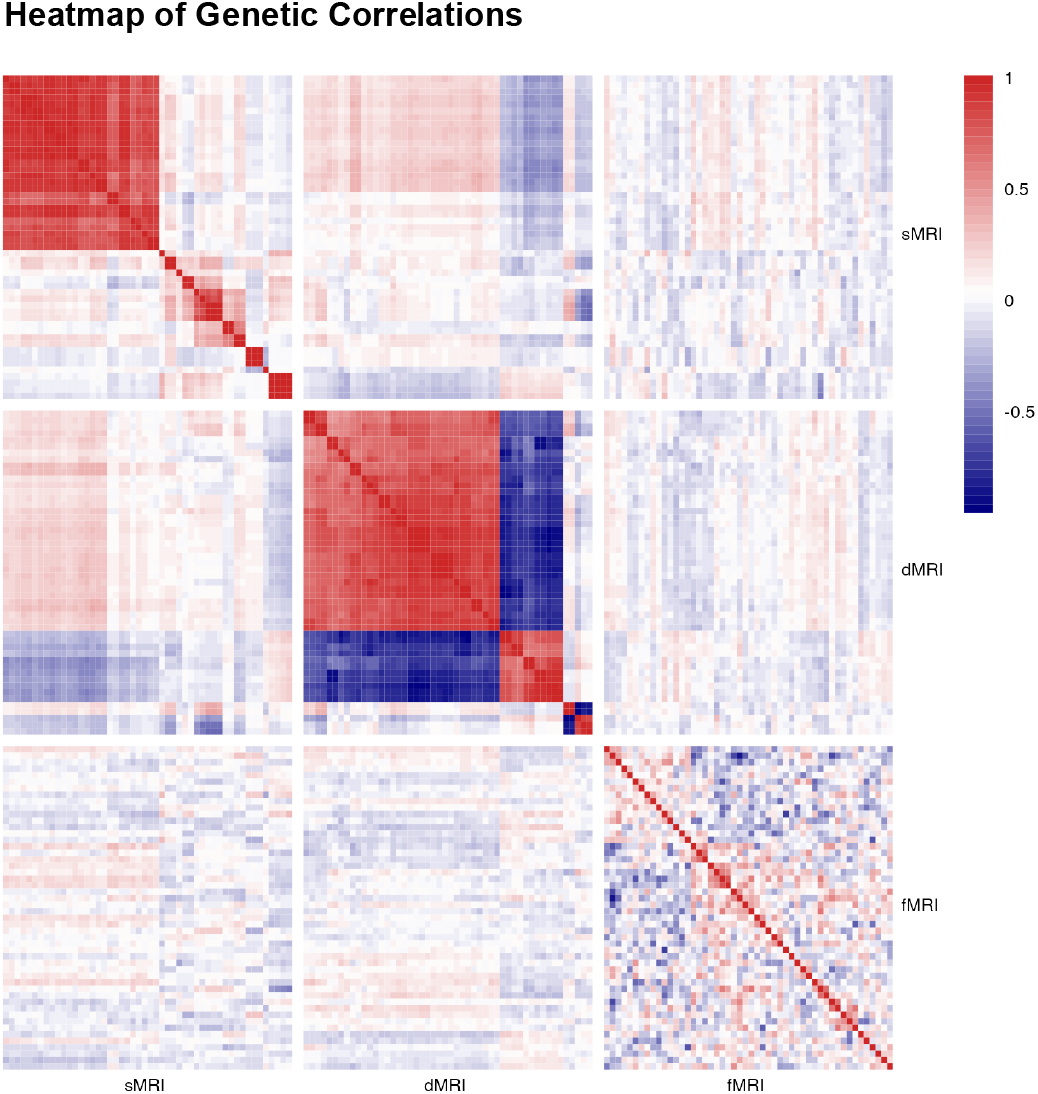
Genetic correlations among the top 50 heritable IDPs (according to Smith et al. 2021) from structural MRI (sMRI), diffusion MRI (dMRI), and functional MRI (fMRI). The correlations were estimated using LDSC, using all SNPs across all chromosomes with summary statistics from UKB IDP GWAS and reference panel from 1000G, with a minor allele frequency (MAF) filter of 1%. IDPs within each modality were re-ordered by hierarchical clustering for better visualization.

##### 2. Multiple testing

Multiple testing becomes an issue when multiple highly correlated IDPs are associated with a gene, increasing the number of comparisons in Stage 2 and making a stringent Bonferroni correction necessary, which can substantially reduce statistical power.

### 2.3 Proposed method: testing modality-specific pathways to AD

To mitigate potential power loss in analyzing multiple IDPs in an IV framework, we propose testing all IDPs in each MRI modality as a unitary entity while controlling for pleiotropic effects from other modalities. Referring back to Figure 1, we largely observed high genetic correlations within each MRI modality, including such high correlated IPDs in MV-IWAS can compromise inference accuracy, which motivated us to test all IDPs at the modality level. Except the high within-modality correlations, we also noted moderate cross-modality correlations, e.g., between sMRI and dMRI IDPs (Figure 1), indicating a need to adjust for cross-modality pleiotropy effects. Beyond statistical considerations, grouping IDPs by modality is also scientifically informative: each MRI modality characterizes distinct aspects of brain structure and function, and therefore, aggregating the brain information in each modality can contribute to a more comprehensive understanding of the genetic mechanisms underlying neurological diseases.

To the end, we specifically aim to answer the question: *how does a gene lead to AD progression through which imaging modality* ? To handle possible bi-directional genetic effects, e.g., bidirectional effect of dMRI IDPs in Figure 1, we propose MV-Modality-IWAS (multivariate modality global testing in IWAS), which bridges the extremes of testing each IDP individually in MV-IWAS (Knutson et al. 2020) and testing all IDPs together through a global testing in IWAS (Xu et al. 2017). By testing all IDPS within each MRI modality, we can assess the genetic regulatory effects of entire sets of imaging features, rather than individual IDPs, which may capture a range of subtle and complex brain processes. Ultimately, this methodology has the potential to uncover novel genetic variants or regions linked to brain structure and function, providing new insights into the underlying mechanisms of AD. A graphical illustration is provided in Figure 2.

**Figure 2.**
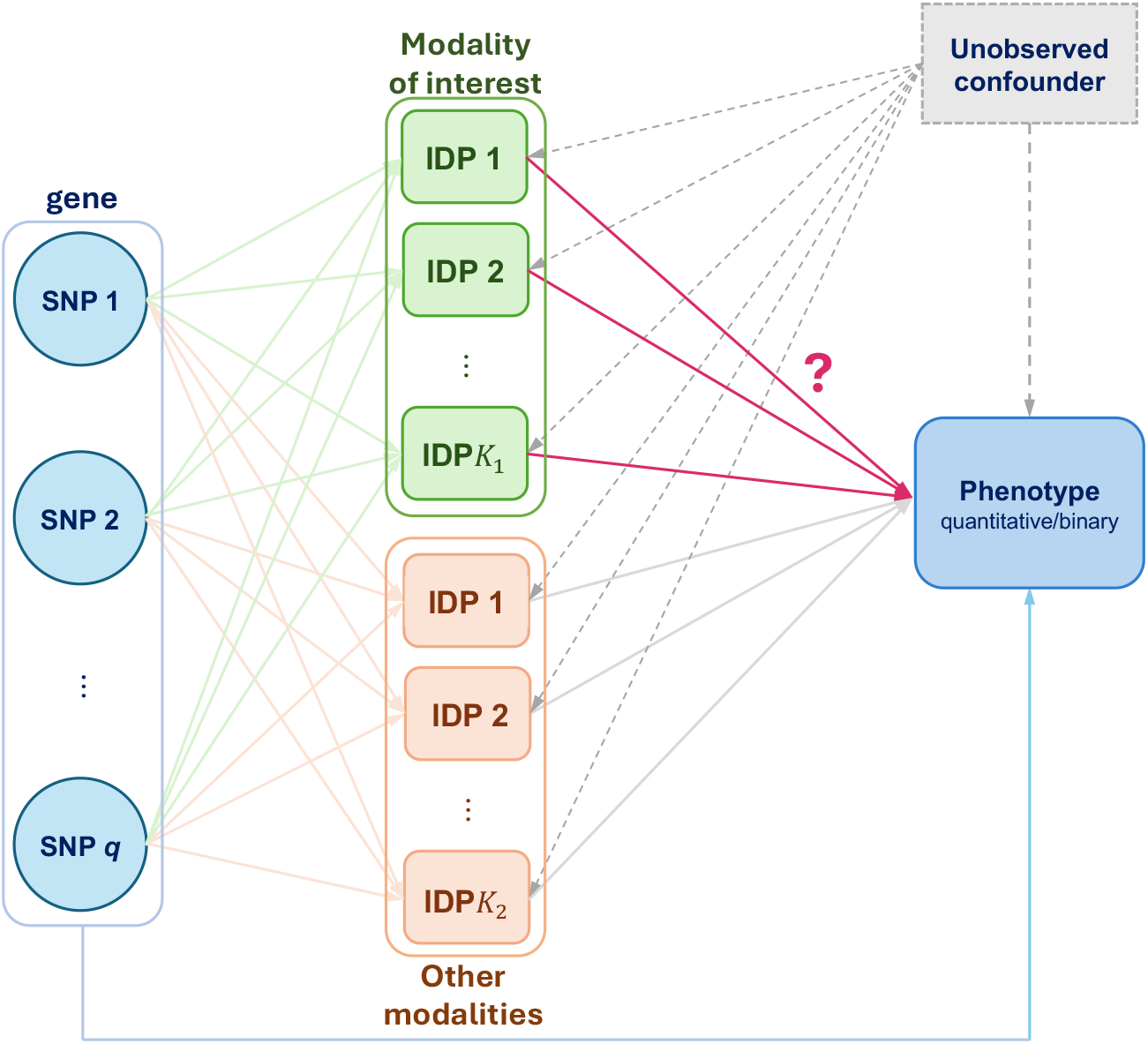
An illustration of a data-generating model for simulation studies, using a directed acyclic graph (DAG) representation. Different colors in IDP denote different imaging modalities, where the green one is the modality of our interest.

Our Stage 2 model consider the testing 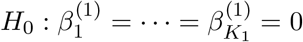 for each gene from the following working model:

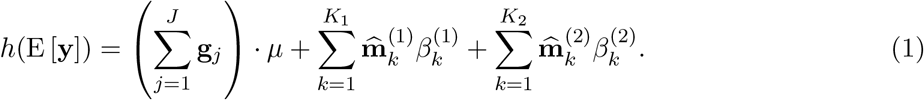

Equation (1) provides an interpretable decomposition of genetic pathways to phenotype: (i) the direct effect (adjusted by *µ*), and (ii) genetically regulated effects (modeled by 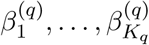) through the *q*th imaging modality. With the assumed model, we test if the specified modality contributes to the phenotype **y**, i.e., test the existence of casual modality-specific genetic pathways.

We propose to test the genetically regulated effects of a given modality using score-based testing while adjusting genetically regulated effects from other modalities, as well as residual genetic direct effects, as fixed effects, with the test statistic *T*,

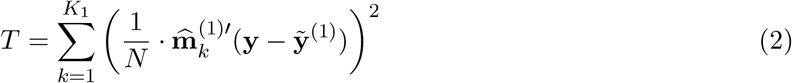

where 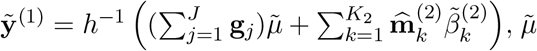 and 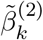 s are estimated under the null hypothesis. Under the null hypothesis, *T* asymptotically follows a mixture 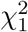 distribution, which is approximated by a single non-central chi-square distribution with matching moments (Davies 1980). In the proposed modality-level testing, the fixed effects of genetically regulated effects from other modalities are adjusted, similarly to the format of the Sequence Kernel Association Test (SKAT) (Wu et al. 2011). The proposed modality-level testing is motivated here well because (i) each IDP contributes only a small proportion to the progression of AD and (ii) the directions (sign) of the effects are expected to be non-uniform. In this scenario, different IDPs may drive AD in varying effect directions and magnitudes, in which SKAT is known to be powerful in this context.

The proposed test is an extension of several previous methods including UV-IWAS and multivariate model in MV-IWAS. If only one IDP in the modality being tested is included, it is equivalent to UV-IWAS. However, the proposed test differs from UV-IWAS in that it adjusts for other modalities, which could lead to horizontal pleiotropy. If the modality has one feature only, the proposed test is also equivalent to MV-IWAS or MV-IWAS-Egger. Our proposed method can be viewed as an extension of previous methods but can handle the challenges of testing genetically regulated effects of imaging features in the modality of interest while adjusting for genetic direct effects and effects from other modalities.

### 2.4 Implementation using GWAS summary statistics

Our method can be implemented using reference GWAS summary statistics, making it more useful in practice. For simplicity in notations, we define **G** = [**g**_1_, …, **g**_*J*_] and let 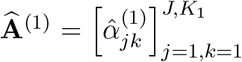 be a coefficient matrix for the modality (of the interest) from the Stage 1 working model for *j* = 1, …, *J* and *k* = 1, …, *K*_1_. We define 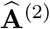 for all other modalities, including direct effects.

Then the Stage 2 model is summarized by testing 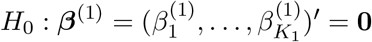 in

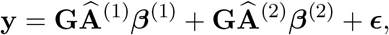

and the test statistic in Equation (2) is equivalent to

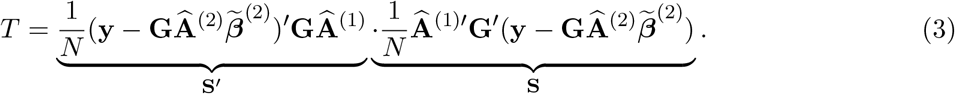

Here,

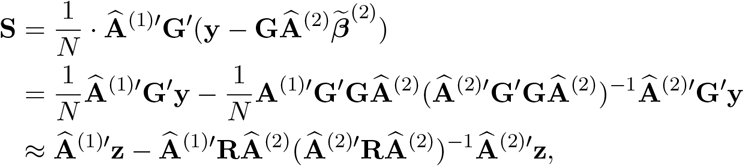

where **z** = (*z*_1_, …, *z*_*J*_) = **G**^*′*^**y***/*(*N* − 1) is the vector of z-scores of the GWAS between the phenotype and a gene, and **R** = **G**^*′*^**G***/*(*N* − 1) is the pairwise Pearson correlation matrix of SNPs in the gene, following Kwak and Pan (2016). Note that **R** could also be approximated by a reference panel (e.g., 1000 Genomes Project (Consortium et al. 2010)). Similarly, we get

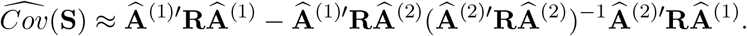

Since **S** follows a multivariate normal distribution with the zero mean under the null hypothesis, it is sufficient to use plug-in estimate 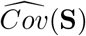 to derive the null distribution of *T* (mixture 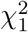). We use the CompQuadForm R package to obtain the *p* value when *K*_1_ *>* 5 and use the Monte Carlo method to obtain the *p* value otherwise to improve suboptimal Davies approximation methods to the mixture 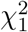 distribution of the test statistic.

We note that our summary statistics approach implicitly assumes that the phenotype of interest is continuous, which is violated when working with binary disease status. However, such ‘linearization’ is a well-accepted method when the effect sizes of the covariates are tiny, which is also the case when evaluating the effects of imputed IDPs to AD (Xue et al. 2020; Knutson et al. 2020). To evaluate the robustness, our simulation studies in Section 3 are implemented by the summary statistics approach in this section, while the logistic model is used. The R functions to implement the proposed summary statistics approach is available at https://github.com/junjypark/MV_VC_IWAS.

## 3. Simulation studies

### 3.1 Simulation designs

We conducted simulation studies to evaluate Type 1 error rates and statistical power. We generated *n* = 2000 subjects and used *J* = 58 SNPs with pairwise genetic correlations obtained from a gene in chromosome 16. The SNP data was generated by using the rmvbin function from the bindata R package.

Two imaging modalities were generated (*q* = 1, 2), each with 10 features. A graphical model in Figure 2 was used to generate simulated imaging features and binary phenotypes. In our simulations, all SNPs were assumed to be associated with all imaging features. There is an unobserved confounding variable **u** which is used to generate imaging features and the phenotype (but not used to compute *p* values). Therefore, the imaging features were generated by

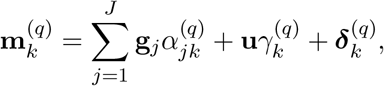

where 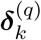 is the residual noise. The binary phenotype was generated via (i) effects from imaging features, (ii) direct effects from genotypes, and (iii) effects from the unobserved confounding variable. Specifically,

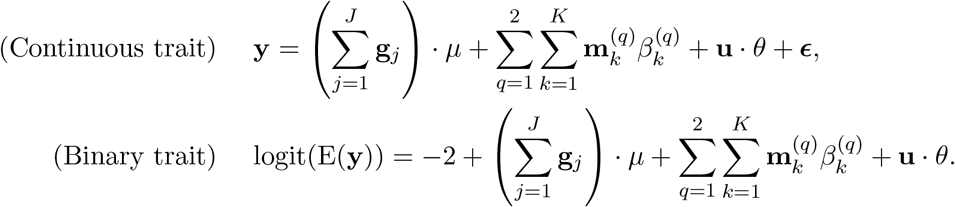

The parameters in our simulation studies were chosen to reflect gene-level effects on imaging features and phenotypes. Specifically, we used 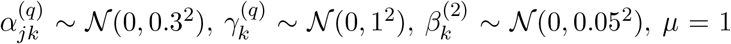 and *θ* = 1. The variance of noise of the IDPs 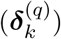 and the continuous trait were set to be 5^2^ and 4^2^, respectively. In both continuous and binary traits, the average *R*^2^ for each of (i) genotype -phenotype and (ii) genotype - IDP was set to be no larger than 0.10 on average under the null model. The average *R*^2^ for IDP - phenotype was also set to be no larger than 0.10 in the continuous trait setting. Also, in binary traits, the prevalence of AD was approximately 18%.

For the power analysis, we consider two different scenarios. In **Scenario 1**, all *q* = 10 IDPs in the first imaging modality causes the trait and 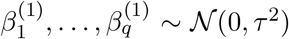 where *τ* controls the degree of signals. In **Scenario 2**, only the first IDP was causal and the remaining IDPs were not (i.e., 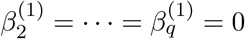.

Denoting our method as **Method 1 (Proposed)**, we compare our methods to three competitors. **Method 2 (IDP-specific)** is the univariate version of the proposed method, where the pleiotropic effect of the other modalities are adjusted but each feature of the modality of interest is tested separately. The global Type 1 error is controlled by applying Bonferroni correction (i.e., *α/*10 for 10 features in the modality). **Method 3 (Proposed unadjusted)** is our method without adjusting for the pleiotropic effect of the other modalities. **Method 4 (UV-IWAS)** is UV-IWAS (i.e., massive-univariate analysis) without adjusting for the pleiotropic effect, with the Bonferronni correction applied.

Since we implemented our data analysis with summary statistics only, we converted each simulated data into necessary summary statistics. For binary traits, GWAS summary statistics were obtained under the true model (e.g., logistic regression), but the assumed stage 2 model was linear regression (Equation (3)). Each simulation was based on 5000 replicated datasets.

### 3.2 Simulation results

We first evaluated the empirical Type 1 error rates of the methods when the first imaging modality does not have a causal effect on the trait. We considered both 5% and 1% as the theoretical rates. The results shown in Figure 3 imply that methods 1 and 2, which consider the pleiotropic effects, controlled Type 1 error rates well, in a slightly conservative manner. Method 2 was more conservative than our methods. It could be explained because each univariate analysis is already conservative (as discussed in Knutson et al. (2020)) and the Bonferroni correction would make it more conservative. Methods 3 and 4 constantly produced inflated Type 1 errors as they do not adjust for pleiotropic effects in their models. Therefore, we only considered methods 1 and 2 in subsequent power evaluations.

**Figure 3.**
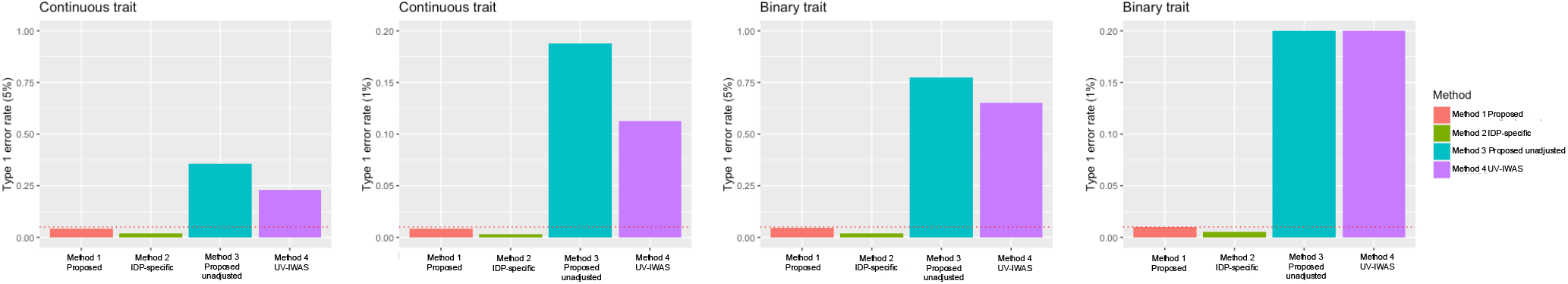
Type 1 error rates of different methods considered in this paper. The red horizontal line denotes the theortical Type 1 error rates. The proposed method controls Type 1 error rate at the nominal rates of 5% and 1% in both continuous and binary traits. Method 3 and 4 suffered from inflated Type 1 error rates as they do not consider pleiotropic effects. For visualizations, the y-axis of the plot was truncated at 0.2 when 1% Type 1 error is applied.

The summaries of power evaluations are provided in Figure 4. We considered Type 1 error rate of 5% when computing power in each scenario. The results were qualitatively similar for both continuous and binary traits. As expected, when all IDPs in the modality of interest has causal effects on the phenotype (“dense signal”), our method achieved higher power than Method 2, and the power increased as the degree of signal increased. Also, since we only used 10 IDPs from each modality, it is expected that the differences between the methods would be more prominent when signals are dense and the number of IDPs increases. When only one IDP had a causal effect on the phenotype, then the power between the two methods were similar. Although the latter result is unexpected because it is a setting that the min*p* method is expected to perform better, it might be explained by the strict Bonferroni correction applied in Method 2 (which was shown in the Type 1 error simulation).

**Figure 4.**
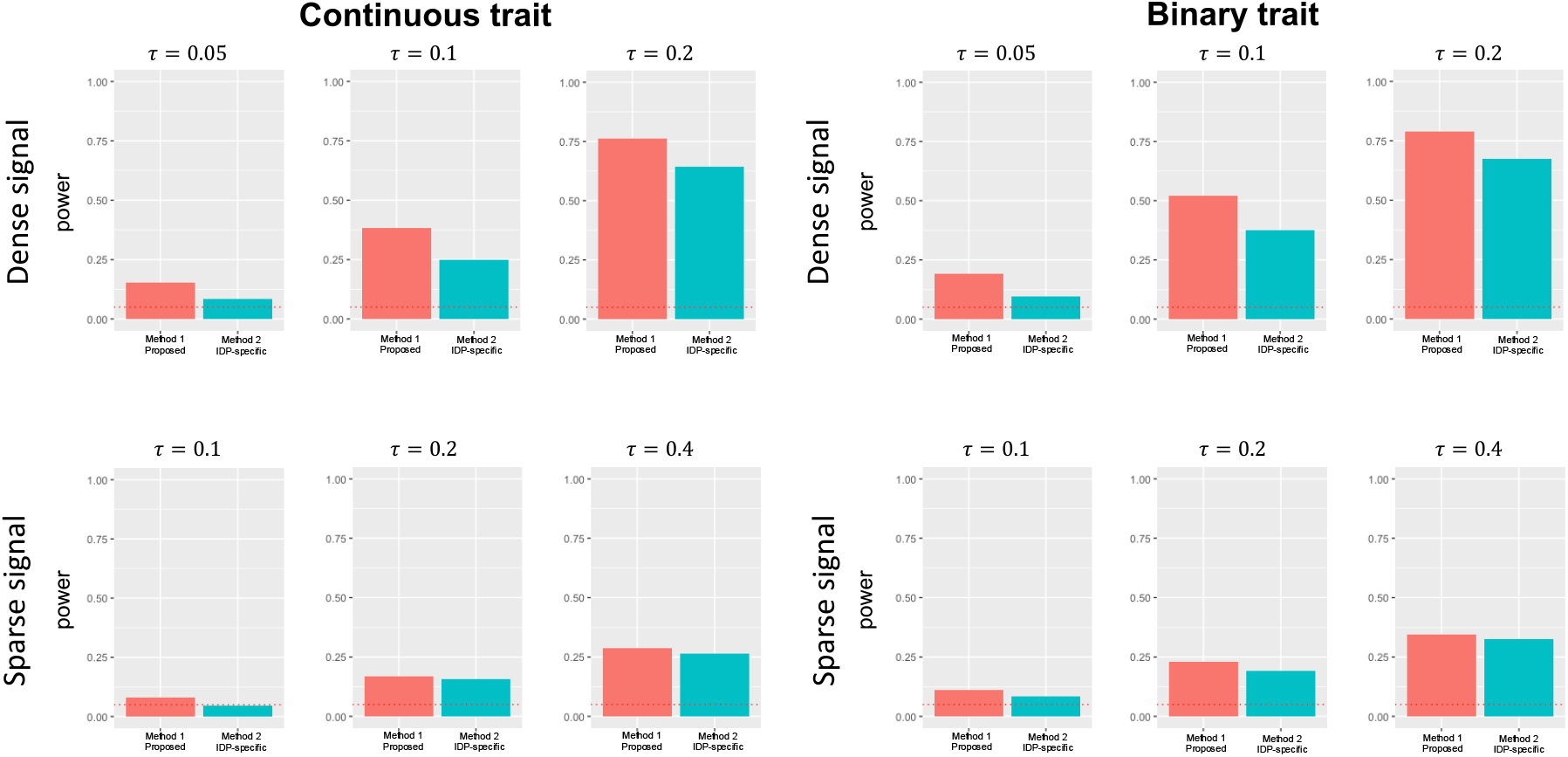
Summary of power analysis. *τ* ^2^ specifies the degree of signals where IDPs in the first modality causes the phenotype. ‘Dense signal’ refers to the case where all IDPs in the first modality causes the hpehnotype, and ‘sparse signal’ refers to the case where only one of the IDPs in the first modality is causal.

## 4 Data analysis

### 4.1 Data availability

#### 4.1.1. UK Biobank: GWAS IDP summary statistics

We analyzed 1436 T1-weighted sMRI features and 690 dMRI features (the IDs and descriptions of these features are available in the supplementary materials) and 210 resting-state functional connectivity metrics from the rfMRI full 25 *×* 25 correlation matrix (data field: 25750). As of 2020, UK Biobank collected approximately 40,000 subjects whose genotype and brain imaging data are both available. Among them, the GWAS summary statistics for each IDP (after adjusting for age, sex, 40 genetic principal components, scanning sites, head size, etc) is provided by Oxford Brain Imaging Genetics Server - BIG40 (https://open.win.ox.ac.uk/ukbiobank/big40/, led by Dr. Lloyd T. Elliott and Dr. Stephen Smith) (Elliott et al. 2018; Smith et al. 2021). The sMRI IDPs are categorized to regional and tissue volume (636 IDPs), cortical area (372 IDPs), cortical thickness (306 IDPs), cortical grey-white contrast (70 IDPs), regional and tissue intensity (41 IDPs). Also, the dMRI IDPs are categorized to WM tract FA (75 IDPs), WM tract MO (75 IDPs), WM tract diffusivity (300 IDPs), WM tract ICVF (75 IDPs), WM tract OD (75 IDPs), WM tract ISOVF (75 IDPs). Genotypes were imputed using the Haplotype Reference Consortium (HRC) as a reference panel (Bycroft et al. 2018).

We used the processing guidelines provided by Knutson et al. (2020) to choose SNPs to be used in our analysis. We used GWAS summary statistics on 2,310 heritable IDPs from 9,707 participants following (Smith et al. 2021), and used 1000G as reference genome for LD clumping. We employed a clumping radius of 1M and set an *r*^2^ cutoff value of 0.5, as recommended by Privé et al. 2019. This approach allowed us to remove loci with high linkage disequilibrium (LD) and keep the most significant SNP in each clump window based on the IDP GWAS results. When dealing with SNPs for a specific gene that are identified from different IDP GWAS clumps, we merged the SNP sets and treated them as a single SNP set for the gene. We refer to https://github.com/kathalexknuts/MVIWAS for more detailed procedures. Then, in our analysis, we only considered variants whose missing rate is less than 1%, and imputed all missing values with median values for each variant. Following Gusev et al. (2016), we considered 1Mb window around each gene to collect SNPs. Then, for each gene, we fitted a general linear model (GLM) to each pre-adjusted imaging feature to obtain weights. Following implications from Knutson et al. (2020), we only considered gene-IDP pairs whose *p* values from the *F* test are less than 5 *×* 10^−5^.

After applying LD clumping, the marginal GWAS vector for each gene was converted to regression coefficients for SNPs → IDP (i.e.,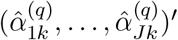) by first transforming the marginal (univariate) GWAS *z* score with 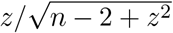(marginal regression coefficient) and multiplying it with the inverse of **R** obtained from 1000 Genomes project.

#### 4.1.2 UK Biobank: GWAS AD summary statistics

In UK Biobank, various health status, including AD, have been phenotyped by ICD 10 codes (data field: 41270). The GWAS summary statistics for AD is provided by the Neale lab (https://www.nealelab.is/uk-biobank), where genotypes are imputed by HRC, UK10K, and 1000 Genomes. The summary statistics were obtained after adjusting for (inferred) sex, age, age^2^, sex*×*age, sex*×*age^2^, and 20 genetic principal components. Among 361194 genotyped subjects, there were approximately 4000 subjects diagnosed with AD.

#### 4.1.3 IGAP: GWAS AD summary statistics

International Genomics of Alzheimer’s Project (IGAP) (Lambert et al. 2013) is a large two-stage study based upon genome-wide association studies (GWAS) on individuals of European ancestry. In stage 1, IGAP used genotyped and imputed data on 7,055,881 SNPs to meta-analyse four previously-published GWAS datasets consisting of 17,008 Alzheimer’s disease cases and 37,154 controls (The European Alzheimer’s disease Initiative – EADI the Alzheimer Disease Genetics Consortium – ADGC The Cohorts for Heart and Aging Research in Genomic Epidemiology consortium – CHARGE The Genetic and Environmental Risk in AD consortium – GERAD). In stage 2, 11,632 SNPs were genotyped and tested for association in an independent set of 8,572 Alzheimer’s disease cases and 11,312 controls. Finally, a meta-analysis was performed combining results from stages 1 and 2. GWAS summary statistics for gene-AD associations were obtained from https://www.niagads.org/datasets/ng00036 based on 17,008 AD cases and 37,154 healthy controls, after adjusting for sex, age, and (at least) four genetic principal components (Lambert et al. 2013).

### 4.2. Analysis results

#### 4.2.1. IGAP: instrumental genes and causal IDPs using IGAP GWAS AD summary statistics

As shown in Figure 5, the instrument genes identified by MV-Modality-IWAS were found in chromosomes 8 (4 genes, 9 genes) and 19 (39 genes, 38 genes) for sMRI and dMRI, respectively, but none was found in fMRI, at the Bonferroni-adjusted significance level of 0.05. Among the genes on chromosome 8, 3 instrumental genes are shared between dMRI and sMRI, 37 instrumental genes are shared between dMRI and sMRI. No instrumental genes were identified for fMRI is as expected, as the fMRI Independent Component Analysis (ICA) was performed at 25 dimensions, summarizing major patterns of brain activity. This approach of data summarizing may results in low (*<* 5%) IDP heritability explained by genotype (Smith et al. 2021), which affects the Stage 1 model and subsequently then leaded to insignificant results in the Stage 2 testing.

**Figure 5.**
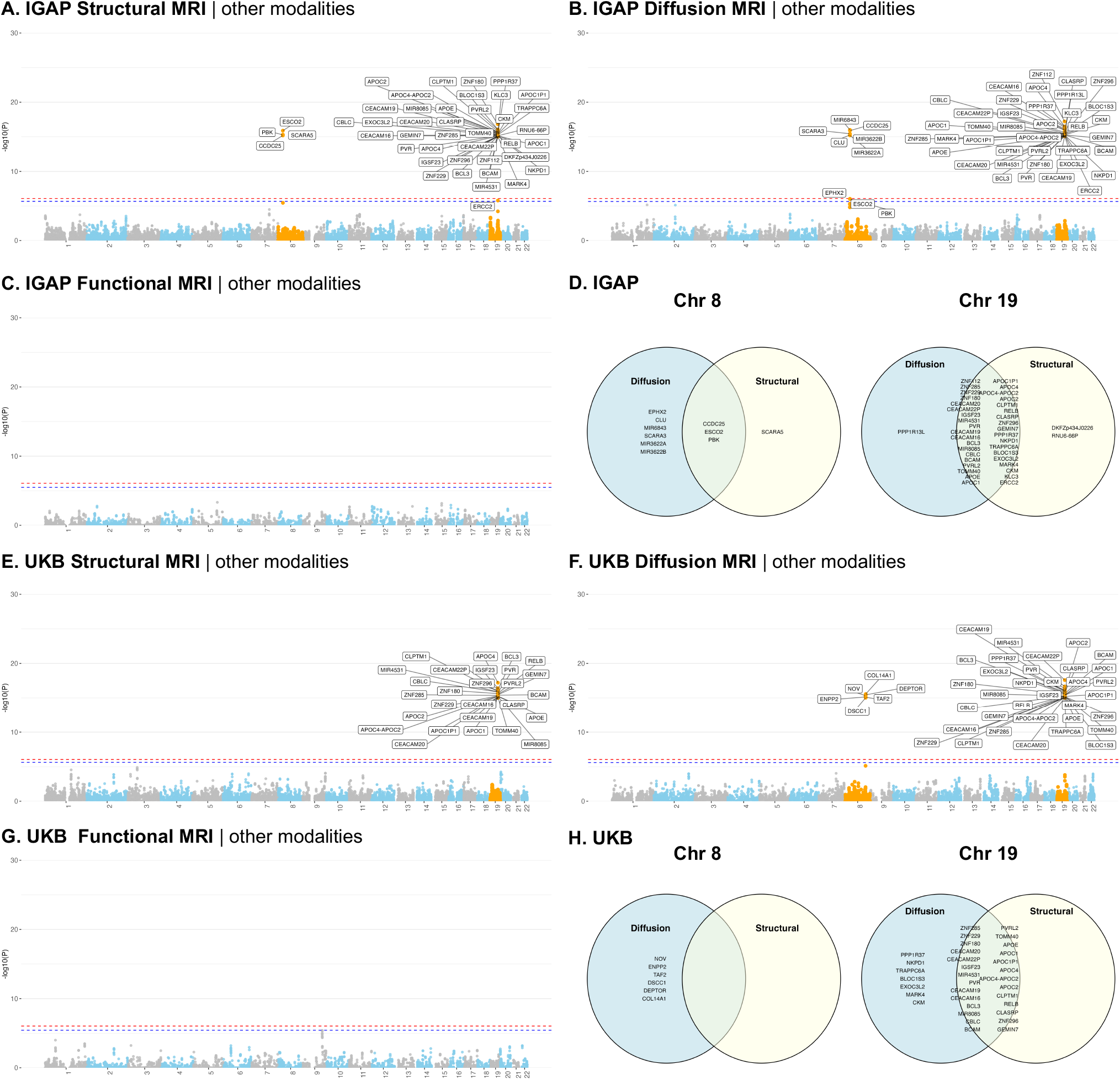
IGAP GWAS AD summary statistics results. Panels A - C show *p*-values for testing causality of each modality when adjusting for pleiotropic effects of other modalities and using each gene as instrumental variables. Panel D shows Venn diagram of the statistically significant genes identified in A - C, summarized by chromosomes 8 and 19. The analysis was performed at a Bonferroni-adjusted significance level, red dashed lines in panel A, B and C represents the global significant threshold i.e., 0.05*/*60, 258; ed dashed lines in panel A, B and C represents the modality-specific significant threshold i.e., 0.05*/*21, 896, 0.05*/*23, 214 and 0.05*/*15, 148, respectively. Panels E - H show the results when using UK Biobank AD GWAS summary statistics. Still, UKB analysis was performed at a Bonferroni-adjusted significance level, red dashed lines in panel E, F and G represents the global significant threshold 0.05*/*54, 082; blue dashed lines in panel E, F and G represents the modality-specific significant threshold i.e., 0.05*/*20, 937, 0.05*/*19, 662 and 0.05*/*13, 483, respectively.

For dMRI, MV-Modality-IWAS identified 71 unique causal IDPs with instrumental genes in chromosome 8 and 19, where 44 IDPs fall in the category of weighted-mean (WM) tract diffusivity, 6 IDPs were WM tract fractional anisotropy (FA), 15 IDPs falls in the category of WM tract intra-cellular volume fraction (ICVF), 3 IDPs fall in the category of WM tract isotropic or free water volume fraction (ISOVF), and 3 IDP falls in the category of WM tract orientation dispersion index (OD).

For sMRI, MV-Modality-IWAS identified 135 unique causal IDPs with instrumental genes in chromosome 8 and 19. Among these, 31 IDP was cortical area, 4 IDPs were cortical grey-white contrasts, 54 IDPs were cortical thickness, 4 IDP were regional and tissue intensities, and 42 IDPs were regional and tissue volume.

#### 4.2.2 UKB: instrumental genes and causal IDPs using UKB GWAS AD summary statistics

With UKB summary statistics, similar to using IGAP summary statistics, we found instrument genes in chromosomes 8 (0 gene, 6 genes) and 19 (27 genes, 34 genes) for sMRI and dMRIs, respectively. However, using UKB summary statistics, fewer instrumental genes were identified overall, particularly for sMRI where no instrumental genes were found on chromosome 8, compared to 4 genes identified using IGAP summary statistics. This discrepancy may be due to the median age difference between the datasets: the median age in UKB is 56.5 years, whereas IGAP considered late-onset AD cases with onset age *>* 65, suggesting potentially stronger signals in IGAP with a larger number of cases used in generating its GWAS summary statistics. Additionally, It is also noteworthy that all instrumental genes identified on chromosome 19 for both dMRI and sMRI using UKB summary statistics are also identified using IGAP summary statistics. However, none of the 6 genes found by UKB for dMRI on chromosome 8 are identified using IGAP summary statistics.

With instrumental genes in chromosome 8 and 19 MV-Modality-IWAS identified 71 unique causal dMRI IDPs for the UKB dataset. Among these 71 unique dMRI IDPs, 36 IDPs fall in the category of WM tract diffusivity, 2 IDPs in WM tract FA, 21 IDPs fall in the category of WM tract ICVF, 6 IDPs fall in the category of WM tract ISOVF, 1 IDP falls in the category of WM tract mean orientation (MO), and 5 IDPs fall in the category of WM tract OD. All causal IDP categories found in IGAP summary statistics were also identified with UKB, with the one additional category: WM tract MO. Additionally, a similar pattern was observed in both IGAP and UKB causal IDP results, where the majority of causal IDPs fell into the categories of WM tract diffusivity and WM tract ICVF.

With instrumental genes in chromosome 19, MV-Modality-IWAS identified 35 unique causal sMRI IDPs from the UKB dataset. Among these, 8 IDPs fall in the category of cortical area, 9 IDPs fall in the category of cortical thickness, 1 IDP falls in the category of regional and tissue intensity, and 17 IDPs fall in the category of regional and tissue volume. The causal IDP categories identified by UKB summary statistics are similar to those of IGAP, but with significantly fewer causal IDPs across all categories. Notably, no causal IDPs were found in cortical grey-white contrasts with UKB data.

Taken altogether, while IGAP and UKB GWAS summary statistics differ by many factors including preprocessing pipelines, the number of samples, the proportion of AD cases, age range, and others, MV-Modality-IWAS identified genes in chromosomes 8 and 19 for sMRI and dMRI and no genes for fMRI in both datasets. These results show potential utility of the proposed method, warranting further investigations.

### 4.3. Comparisons

To test alternative methods of identifying causal IDPs, we compared our proposed approach with two additional strategies. With these comparisons we aim to explore different underlying model assumptions and potential biases in identifying causal MRI-AD relationships.

#### 4.3.1. No adjustment for horizontal pleiotropy effects

In this comparison, we tested the same hypothesis without adjusting for pleiotropic effects from other modalities. The summaries of *p* values are provided in Figure 6. In sMRI analysis, when compared to MV-Modality-IWAS, ignoring pleiotropy found 9 additional genes from chromosome 2, but did not detect the loci in chromosome 8 in IGAP. In UKB, however, no instrumental genes are detected, which yielded heterogeneity in results. In dMRI analysis, ignoring pleiotropy found 8 genes from chromosome 1 and 9 genes in chromosome 2 but did not detect the loci in chromosome 8 in IGAP. In UKB, the loci in chromosomes 1 and 2 did not show any significance, but there was a loci (with 3 genes) in chromosome 22. In fMRI analysis, while UKB did not show any significant gene (which agreed with MV-Modality-IWAS results), IGAP detected 1 instrumental gene on chromosome 1 and 35 instrumental genes on chromosome 19. These noticeable differences in results indicate potential false positive genes from the lens of causal inference.

**Figure 6.**
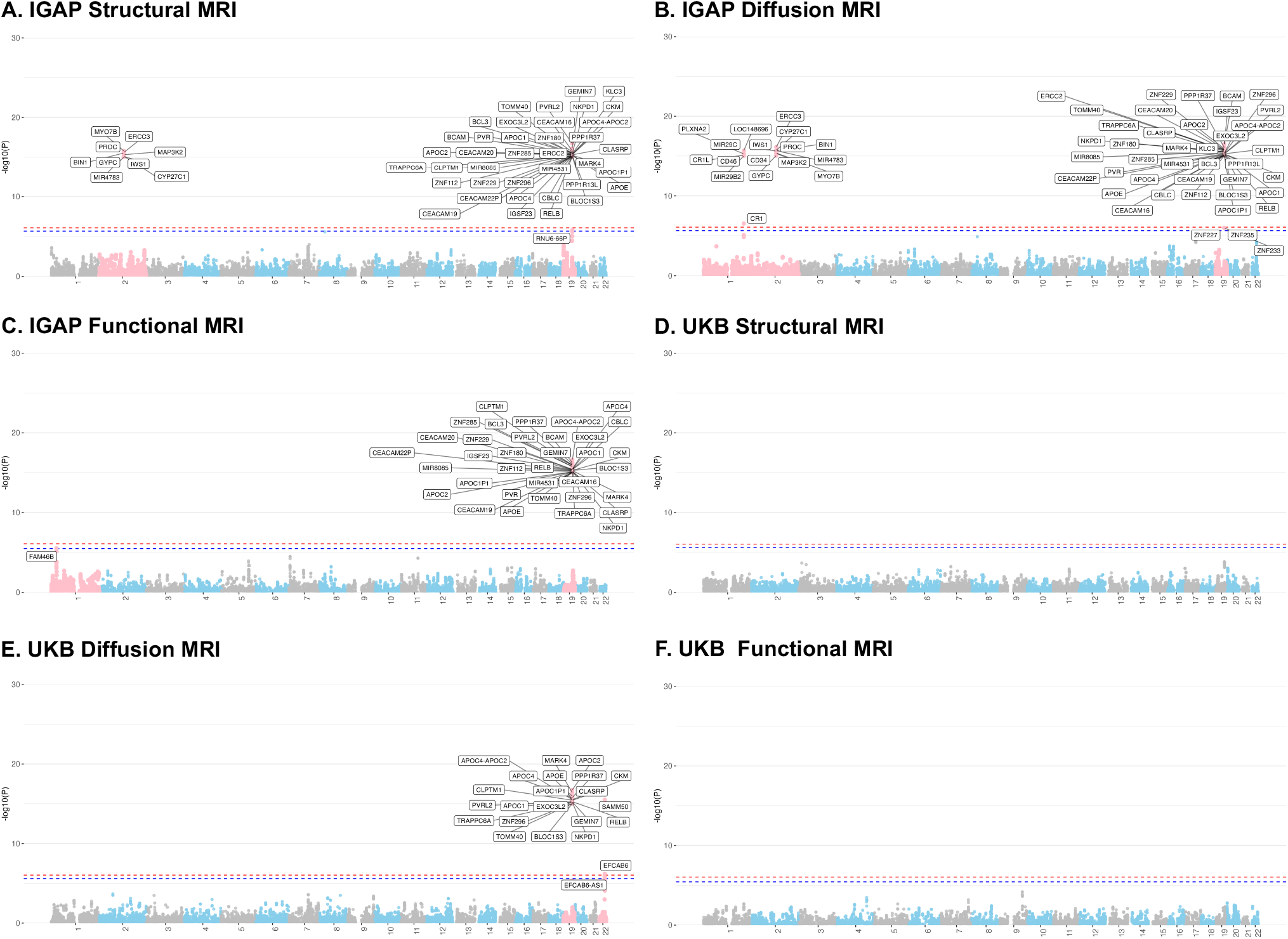
IGAP GWAS AD summary statistics results. Panels A - C show *p*-values for testing causality of each modality when when no pleiotropy effects are adjusted and using each gene as instrumental variables. The analysis was performed at a Bonferroni-adjusted as describied in Figure 4. Panels D - F show the results when using UK Biobank AD GWAS summary statistics and no pleiotropy effects are adjusted. UKB analysis was performed at a Bonferroni-adjusted significance level as describied in Figure 4.

#### 4.3.2. MV-IWAS

In this comparison, we test each IDP individually while adjusting for all other IDPs within the same modality as well as IDPs from other modalities. To control for multiple comparisons in modality level testing, a Bonferroni correction was applied based on the number of tests conducted within each modality. Specifically, there were 699,007 tests in total being conducted in IGAP GWAS and 615,315 tests in total conducted in UKB GWAS. We did not find any statistical significance in both analyses. It’s also important to note that our approach to performing MV-IWAS differs from the Knutson et al. (2020): (1) in the original approach, all SNPs in all chromosomes are applied in the Stage 1 model for impute each IDP, whereas we only used the SNPs within each gene being tested; (2) in the original MV-IWAS, only 14 IDPs were tested, so the Bonferroni correction had a much milder influence compared to our individual IDP testing, which involves a larger number of tests.

These results support our proposal of testing MRI modalities as a whole unit. While testing each IDP individually offers better interpretation regarding causality in identifying causal IDPs, it is too restrictive that no statistically significant results were found using both IGAP and UKB data after applying Bonferroni correction. Analyzing each IDP individually necessitates a much stronger signal to achieve significant findings, which underscores the challenge of power loss with this approach.

## 5. Discussion

In this study, we were motivated by the large amount of MRI data available in the UK Biobank and the power loss associated with analyzing a large number of individual IDPs using existing methods such as MV-IWAS. To address this issue, we proposed an association test for the genetically regulated effects of a given MRI modality (sMRI, dMRI or fMRI) on the progression of Alzheimer’s disease with high power. Our method views each MRI modality as a unitary entity and tests the genetically regulated effects of the modality as a whole. Finally, since each IDP contributes only a small proportion to the progression of AD, we propose a variance component test to test if the overall genetically regulated effects of one modality contribute to the progression of AD while adjusting for the effects of other modalities as well as the residual genetic direct effects.

The proposed method, MV-Modality-IWAS, provides a new perspective on how to handle high-dimensional complex imaging data in genetic association studies. Our method significantly reduces the number of tests needed, resulting in higher power and a reduced need for family-wise error correction, especially when dealing with a large number of IDPs. Similar to MV-IWAS, our method adjusts for other genetically regulated effects and residual genetic direct effects, thus providing better control of type I error than UV-IWAS. Furthermore, by collapsing the information in the modality being tested, fewer parameters are being tested, allowing our method to achieve higher power while keeping the type I error low. Overall, our method provides a powerful and efficient approach to analyzing complex imaging data in genetic association studies. However, there is a trade-off between power and causal interpretability. While the proposed method can identify the modality of the genetically regulated effect, we cannot pinpoint the specific IDPs through which the effect is mediated. Therefore, compared to MV-IWAS (or MV-IWAS-Egger), the proposed method can achieve higher power but cannot conclude the causal genetically regulated effect of individual IDPs within the modality.

In general, causal inference on observational data is not without assumptions and limitations, and imaging genetic studies may also be subject to violations on assumptions. First, we only listed brain imaging data as potential exposures. In practice, there could be environmental/behavioral factors or other physiological measures (including -*omics* or other imaging types such as heart MRI (Zhao et al. 2023) and retinal optical coherence tomography (Zhao et al. 2024)) that could serve as potential exposures. In this case, it is expected that our causal argument would be weaker. Second, we considered horizontal pleiotropy in our settings only but not vertical pleiotropy, which might be the case in neuroimaging when a certain modality (e.g., sMRI) leads changes in other modality (e.g., functional MRI). Despite these limitations, our proposed method makes one step closer in addressing high-dimensionality of pleiotropic effects in leveraging IDPs in studying genetic effects on AD. We also note that these are ongoing challenges in causal inference in genetics research where more research is needed to make valid inferences in such cases.

One possible extension is to use a dimension reduction method (e.g., principal component analysis or independent component analysis) to relax issues with multicollinearity when horizontal pleiotropy exists (Mo et al. 2021; Karageorgiou et al. 2023; Patel et al. 2024). It might require determination of dimensions in each modality. It could be a popular method such as principal component analysis or independent component analysis, or a more flexible methods that decompose and factorize multi-modal data into ‘shared and individual’ variations (Lock et al. 2013; Park and Lock 2020; Lock et al. 2022). Although it is not evaluated in this paper because it requires access to individual-level genotype and imaging data, as well as computational costs to train machine learning models for each gene-IDP pair, we believe such approaches would greatly enhance the scope of our method. Especially if there is availability of dimensionality-reduced composite score GWAS summary statistics, and with clear evidence that the genetic variation is captured by the composite scores, the dimensionality-reduced composite scores can be used to replace the IDPs in the analysis.

## Data Availability

The GWAS summary statistics data used in this study is publicly available, with details provided in Section 4.1 of the preprint. Codes used in simulation studies and data analyses are available at https://github.com/junjypark/MV_VC_IWAS.

## Author Contributions

**YT**: Data curation, Formal analysis, Methodology, Software, Validation, Visualization, Writing – original draft. **DF**: Funding acquisition, Supervision, Writing – review & editing. **JG**: Funding acquisition, Supervision, Writing – review & editing. **JYP**: Conceptualization, Formal analysis, Funding acquisition, Methodology, Software, Supervision, Writing – original draft, Writing – review & editing

## Data and Code Availability

The GWAS summary statistics data used in this study is publicly available, with details provided in Section 4.1. Codes used in simulation studies and data analyses are available at https://github.com/junjypark/MV_VC_IWAS.

## Ethics Statement

No individual-level genetic data was accessed or analyzed in this study. Ethical approval for this study was reviewed and granted by the Office of Research Ethics, University of Toronto (protocol number 44674).

## Acknowledgements

We thank the editor, associate editor, and three anonymous reviewers for their valuable suggestions and comments, which improved the quality of the manuscript. We also thank Dr. Lisa Strug and Dr. Lei Sun (University of Toronto) for providing helpful suggestions and comments on this work. We thank Wellcome Centre for Integrative Neuroimaging (WIN/FMRIB), Oxford, UK and Department of Statistics and Actuarial Science, Simon Fraser University, Canada for providing GWAS summary statistics for imaging-driven phenotypes from the UK Biobank cohort through the Oxford Brain Imaging Genetics Server - BIG40. We also thank the Neale lab for providing GWAS summary statistics for Alzheimer’s disease from the UK Biobank cohort, and the International Genomics of Alzheimer’s Project (IGAP) for providing summary results data for these analyses. The investigators within IGAP contributed to the design and implementation of IGAP and/or provided data but did not participate in analysis or writing of this report. IGAP was made possible by the generous participation of the control subjects, the patients, and their families. The i–Select chips was funded by the French National Foundation on Alzheimer’s disease and related disorders. EADI was supported by the LABEX (laboratory of excellence program investment for the future) DISTALZ grant, Inserm, Institut Pasteur de Lille, Université de Lille 2 and the Lille University Hospital. GERAD was supported by the Medical Research Council (Grant no 503480), Alzheimer’s Research UK (Grant no 503176), the Wellcome Trust (Grant no 082604/2/07/Z) and German Federal Ministry of Education and Research (BMBF): Competence Network Dementia (CND) grant no 01GI0102, 01GI0711, 01GI0420. CHARGE was partly supported by the NIH/NIA grant R01 AG033193 and the NIA AG081220 and AGES contract N01–AG–12100, the NHLBI grant R01 HL105756, the Icelandic Heart Association, and the Erasmus Medical Center and Erasmus University. ADGC was supported by the NIH/NIA grants: U01 AG032984, U24 AG021886, U01 AG016976, and the Alzheimer’s Association grant ADGC–10–196728.

This research was supported by the McLaughlin Centre (accelerator grant). DF was supported by The Koerner Family Foundation, The Krembil Foundation, The CAMH Foundation, and the Canadian Institutes of Health Research. JG was partially supported by the Natural Sciences and Engineering Research Council of Canada (RGPIN-2021-03734), and the University of Toronto’s Data Science Institute, and the Connaught Fund. JYP was partially supported by the Natural Sciences and Engineering Research Council of Canada (RGPIN-2022-04831), and the University of Toronto’s Data Science Institute, and the Connaught Fund.

## References

Burgess, Stephen, Verena Zuber, Elsa Valdes-Marquez, Benjamin B Sun, and Jemma C Hopewell (2017). “Mendelian randomization with fine-mapped genetic data: choosing from large numbers of correlated instrumental variables”. In: Genetic Epidemiology 41.8, pp. 714–725. doi: 10.1002/gepi.22077.

Bycroft, Clare, Colin Freeman, Desislava Petkova, Gavin Band, Lloyd T Elliott, Kevin Sharp, Allan Motyer, Damjan Vukcevic, Olivier Delaneau, Jared O’Connell, et al. (2018). “The UK Biobank resource with deep phenotyping and genomic data”. In: Nature 562.7726, pp. 203–209. doi: 10.1038/s41586-018-0579-z.

Chan, Lap Sum, Mykhaylo M Malakhov, and Wei Pan (2024). “A novel multivariable Mendelian randomization framework to disentangle highly correlated exposures with application to metabolomics”. In: The American Journal of Human Genetics. doi: 10.1016/j.ajhg.2024.07.007.

Consortium, 1000 Genomes Project et al. (2010). “A map of human genome variation from population scale sequencing”. In: Nature 467.7319, p. 1061. doi: 10.1038/nature09534.

Davies, Robert B (1980). “The distribution of a linear combination of χ2 random variables”. In: Journal of the Royal Statistical Society Series C: Applied Statistics 29.3, pp. 323–333. doi: 10.2307/2346911.

Duncan, Laramie E, Hanyang Shen, Jacob S Ballon, Kate V Hardy, Douglas L Noordsy, and Douglas F Levinson (2018). “Genetic correlation profile of schizophrenia mirrors epidemiological results and suggests link between polygenic and rare variant (22q11. 2) cases of schizophrenia”. In: Schizophrenia Bulletin 44.6, pp. 1350–1361.

Elliott, Lloyd T, Kevin Sharp, Fidel Alfaro-Almagro, Sinan Shi, Karla L Miller, Gwenäelle Douaud, Jonathan Marchini, and Stephen M Smith (2018). “Genome-wide association studies of brain imaging phenotypes in UK Biobank”. In: Nature 562.7726, pp. 210–216. doi: 10.1038/s41586-018-0571-7.

Gamazon, Eric R, Heather E Wheeler, Kaanan P Shah, Sahar V Mozaffari, Keston Aquino-Michaels, Robert J Carroll, Anne E Eyler, Joshua C Denny, Dan L Nicolae, Nancy J Cox, et al. (2015). “A gene-based association method for mapping traits using reference transcriptome data”. In: Nature Genetics 47.9, pp. 1091–1098. doi: 10.1038/ng.3367.

Gatz, Margaret, Chandra A Reynolds, Laura Fratiglioni, Boo Johansson, James A Mortimer, Stig Berg, Amy Fiske, and Nancy L Pedersen (2006). “Role of genes and environments for explaining Alzheimer disease”. In: Archives of General Psychiatry 63.2, pp. 168–174. doi: 10.1001/archpsyc.63.2.168.

Gusev, Alexander, Arthur Ko, Huwenbo Shi, Gaurav Bhatia, Wonil Chung, Brenda WJH Penninx, Rick Jansen, Eco JC De Geus, Dorret I Boomsma, Fred A Wright, et al. (2016). “Integrative approaches for large-scale transcriptome-wide association studies”. In: Nature genetics 48.3, pp. 245– 252. doi: 10.1038/ng.3506.

Huang, Meiyan, Thomas Nichols, Chao Huang, Yang Yu, Zhaohua Lu, Rebecca C Knickmeyer, Qianjin Feng, Hongtu Zhu, Alzheimer’s Disease Neuroimaging Initiative, et al. (2015). “FVGWAS: Fast voxelwise genome wide association analysis of large-scale imaging genetic data”. In: NeuroImage 118, pp. 613–627. doi: 10.1016/j.neuroimage.2015.05.043.

Karageorgiou, Vasileios, Dipender Gill, Jack Bowden, and Verena Zuber (2023). “Sparse dimensionality reduction approaches in Mendelian randomisation with highly correlated exposures”. In: Elife 12, e80063. doi: 10.7554/eLife.80063.

Knutson, Katherine A, Yangqing Deng, and Wei Pan (2020). “Implicating causal brain imaging endophenotypes in Alzheimer’s disease using multivariable IWAS and GWAS summary data”. In: NeuroImage 223, p. 117347. doi: 10.1016/j.neuroimage.2020.117347.

Kwak, Il-Youp and Wei Pan (2016). “Adaptive gene-and pathway-trait association testing with GWAS summary statistics”. In: Bioinformatics 32.8, pp. 1178–1184. doi: 10.1093/bioinformatics/btv719.

Lambert, Jean-Charles, Carla A Ibrahim-Verbaas, Denise Harold, Adam C Naj, Rebecca Sims, Céline Bellenguez, Gyungah Jun, Anita L DeStefano, Joshua C Bis, Gary W Beecham, et al. (2013). “Meta-analysis of 74,046 individuals identifies 11 new susceptibility loci for Alzheimer’s disease”. In: Nature genetics 45.12, pp. 1452–1458. doi: 10.1038/ng.2802.

Liang, Yanyu, Owen Melia, Timothy J Caroll, Thomas Brettin, Andrew Brown, and Hae Kyung Im (2022). “BrainXcan identifies brain features associated with behavioral and psychiatric traits using large scale genetic and imaging data”. In: medRxiv, pp. 2021–06. doi: 10.1101/2021.06.01.21258159.

Lin, Dongdong, Hongbao Cao, Vince D Calhoun, and Yu-Ping Wang (2014). “Sparse models for correlative and integrative analysis of imaging and genetic data”. In: Journal of Neuroscience Methods 237, pp. 69–78. doi: 10.1016/j.jneumeth.2014.09.001.

Lock, Eric F, Katherine A Hoadley, James Stephen Marron, and Andrew B Nobel (2013). “Joint and individual variation explained (JIVE) for integrated analysis of multiple data types”. In: The Annals of Applied Statistics 7.1, p. 523. doi: 10.1214/12-AOAS597.

Lock, Eric F, Jun Young Park, and Katherine A Hoadley (2022). “Bidimensional linked matrix factorization for pan-omics pan-cancer analysis”. In: The Annals of Applied Statistics 16.1, p. 193. doi: 10.1214/21-AOAS1495.

Mai, Jialin, Mingming Lu, Qianwen Gao, Jingyao Zeng, and Jingfa Xiao (2023). “Transcriptome-wide association studies: recent advances in methods, applications and available databases”. In: Communications Biology 6.1, p. 899. doi: 10.1038/s42003-023-05279-y.

Mo, Chen, Zhenyao Ye, Hongjie Ke, Tong Lu, Travis Canida, Song Liu, Qiong Wu, Zhiwei Zhao, Yizhou Ma, L Elliot Hong, et al. (2021). “A new Mendelian Randomization method to estimate causal effects of multivariable brain imaging exposures”. In: Pacific Symposium on Biocomputing 2022. World Scientific, pp. 73–84. doi: 10.1142/9789811250477_0008.

Park, Jun Young and Eric F Lock (2020). “Integrative factorization of bidimensionally linked matrices”. In: Biometrics 76.1, pp. 61–74. doi: 10.1111/biom.13141.

Patel, Ashish, Dipender Gill, Dmitry Shungin, Christos S Mantzoros, Lotte Bjerre Knudsen, Jack Bowden, and Stephen Burgess (2024). “Robust use of phenotypic heterogeneity at drug target genes for mechanistic insights: Application of cis-multivariable Mendelian randomization to GLP1R gene region”. In: Genetic Epidemiology 48.4, pp. 151–163. doi: 10.1002/gepi.22551.

Privé, Florian, Bjarni J Vilhjálmsson, Hugues Aschard, and Michael GB Blum (2019). “Making the most of clumping and thresholding for polygenic scores”. In: The American Journal of Human Genetics 105.6, pp. 1213–1221. doi: 10.1016/j.ajhg.2019.11.001.

Shen, Li and Paul M Thompson (2019). “Brain imaging genomics: integrated analysis and machine learning”. In: Proceedings of the IEEE 108.1, pp. 125–162. doi: 10.1109/JPROC.2019.2947272.

Smith, Stephen M, Gwenäelle Douaud, Winfield Chen, Taylor Hanayik, Fidel Alfaro-Almagro, Kevin Sharp, and Lloyd T Elliott (2021). “An expanded set of genome-wide association studies of brain imaging phenotypes in UK Biobank”. In: Nature Neuroscience 24.5, pp. 737–745. doi: 10.1038/s41593-021-00826-4.

Tanzi, Rudolph E (2012). “The genetics of Alzheimer disease”. In: Cold Spring Harbor perspectives in medicine 2.10, a006296. doi: 10.1101/cshperspect.a006296.

Taschler, Bernd, Stephen M Smith, and Thomas E Nichols (2022). “Causal inference on neuroimaging data with Mendelian randomisation”. In: NeuroImage 258, p. 119385. doi: 10.1016/j.neuroimage.2022.119385.

Wu, Michael C, Seunggeun Lee, Tianxi Cai, Yun Li, Michael Boehnke, and Xihong Lin (2011). “Rare-variant association testing for sequencing data with the sequence kernel association test”. In: The American Journal of Human Genetics 89.1, pp. 82–93. doi: 10.1016/j.ajhg.2011.05.029.

Xu, Zhiyuan, Chong Wu, Wei Pan, Alzheimer’s Disease Neuroimaging Initiative, et al. (2017). “Imaging-wide association study: integrating imaging endophenotypes in GWAS”. In: NeuroImage 159, pp. 159–169. doi: 10.1016/j.neuroimage.2017.07.036.

Xue, Haoran, Wei Pan, and Alzheimer’s Disease Neuroimaging Initiative (2020). “Some statistical consideration in transcriptome-wide association studies”. In: Genetic Epidemiology 44.3, pp. 221– 232. doi: 10.1002/gepi.22274.

Zhao, Bingxin, Tengfei Li, Zirui Fan, Yue Yang, Juan Shu, Xiaochen Yang, Xifeng Wang, Tianyou Luo, Jiarui Tang, D. Xiong, et al. (2023). “Heart-brain connections: Phenotypic and genetic insights from magnetic resonance images”. In: Science 380.6648, abn6598. doi: 10.1126/science.abn6598.

Zhao, Bingxin, Yujue Li, Zirui Fan, Zhenyi Wu, Juan Shu, Xiaochen Yang, Yilin Yang, Xifeng Wang, Bingxuan Li, Xiyao Wang, et al. (2024). “Eye-brain connections revealed by multimodal retinal and brain imaging genetics”. In: Nature Communications 15.1, p. 6064. doi: 10.1038/s41467-024-50309-w.

